# Functional implications of aging-related lncRNAs for predicting prognosis and immune status in glioma patients

**DOI:** 10.1101/2021.12.26.21268427

**Authors:** Guangying Zhang, Yanyan Li, Na Li, Liangfang Shen, Zhanzhan Li

## Abstract

Glioma, is the most prevalent intracranial tumor with high recurrence and mortality rate. Long noncoding RNAs (lncRNAs) play a critical role in the occurrence and progression of tumors as well as in aging regulation. Our study aimed to establish a new glioma prognosis model by integrating aging-related lncRNAs expression profiles and clinical parameters in glioma patients from the Chinese Glioma Genome Atlas (CGGA) and the Cancer Genome Atlas (TCGA) datasets. The Pearson correlation analysis (|R|> 0.6, P<0.001) was performed to explore the aging-related lncRNAs, and univariate cox tregresion and least absolute shrinkage and selection operator (LASSO) regression were used to screening prognostic signature in glioma patients. Based on the fifteen lncRNAs, we can divide glioma patients into three subtypes, and developed a prognostic model. Kaplan-Meier survival curve analysis showed that low-risk patients had longer survival time than high-risk group. Principal component analysis indicated that aging-related lncRNAs signature had a clear distinction between high- and low-risk groups. We also found that fifteen target lncRNAs were closely correlated with 119 genes by establishing a co-expression network. In addition, Kyoto Encyclopedia of Genes and Genomes (KEGG) analysis displayed different function and pathways enrichment in high-and low-risk groups. The different missense mutations were observed in two groups, and the most frequent variant types were single nucleotide polymorphism (SNP). This study demonstrated that the novel aging-related lncRNAs signature had an important prognosis prediction and may contribute to individual treatment for glioma.

## Introduction

Glioma is a highly heterogeneous tumor in central nervous system with high recurrence and mortality rate, especially glioblastoma. The median survival time of WHO III grade glioma is approximately 3 years, whereas WHO IV grade glioma has a grave prognosis of less than 15 months[1]. The conventional treatment for gliomas is surgical resection, radiotherapy and temozolomide (TMZ) chemotherapy. The drug treatment of glioma remains challenging due to the unique structure of blood-brain barrier, which prevents numerous antitumor drugs into the brain[2, 3]. Despite various cancer therapies have been applied over the past decades, the prognosis of glioma patients remains dismal. And the prognosis of glioma patients with the same grade is inconsistent. Recently, it is proved that many mutations could assess risk factor and predict prognosis of glioma, such as isocitrate dehydrogenase (IDH) mutation and 1p19q co-deletion, which present relatively favorable survival[4]. But some of these indicators are not very comprehensive to predict the prognosis of all grade glioma patients, for example, IDH (since nearly 80% of patients with low-grade glioma have IDH mutations). Therefore, a robust prognostic model is urgently needed to predict survival of patients with the malignant growth and highly relapse rate of glioma.

Long non-coding RNAs (lncRNAs) are more than 200 nucleotides in length, which do not encode protein and involve in post-transcriptional modulation and gene translation[5]. It is recorded that the diversity of lncRNAs is implicated in many biological functions, such as epigenetic regulation, tumor microenvironment and cell apoptosis[6]. Some scholars have revealed that overexpression of lncRNA AGAP2-AS1 enhanced breast tumor growth and trastuzumab resistance[7]. The high expression of HOTAIR also promotes proliferation of breast tumor cells and drug resistance to tamoxifen[8]. Luo et al. have found that the depletion of lncRNA AGAP2-AS1 depressed proliferation and invasion, and induced apoptosis in U251 cells[9]. Other study has shown that overexpression of ATB is predicts a poor prognosis in colorectal cancer[10]. With further research on lncRNAs, accumulating researches have been identified lncRNAs as critical roles in regulating cell proliferation, invasion, apoptosis and drug resistance in a variety of tumors[11, 12]. Meanwhile, the vital roles of lncRNAs in degenerative disease of the central nervous system are becoming evident[13]. LncRNAs also promote tumor immune evasion, for example, NKILA escape immunological destruction by sensitizing T cells and inhibit NF-κB activity[14].

As is known to us, neurodegenerative disorders and cancer are age-related diseases. Recent researches have demonstrated that lncRNA expression profiles are influencing aging. An accumulation of studies has suggested that aging is a significant risk factor for the development of cancer[15, 16]. However, the role of aging-related lncRNAs in gliomas has not been fully elucidated. In the present study, we integrated gene matrix and clinical parameters from CGGA and TCGA database. Fifteen lncRNAs related to aging are screened by cox regression analyses. The aging-related lncRNAs prognosis model can predict the prognosis of glioma patients as a potential prognostic indicator, which may provide crucial implication in clinical targeted therapy.

## Materials and methods

### Data source

The complete RNA-sequencing data and corresponding clinical features of glioma patients were obtained from the Chinese Glioma Genome Atlas (CGGA) and The Cancer Genome Atlas (TCGA; https://cancergenome.nih.gov/). The lncRNA and protein-coding genes were classified according to gene annotation in GENCODE project (https://www.gencodegenes.org/) form our downloaded raw readings and fragments per kilo-base of transcript per million data. Detailed clinicopathological information, including age, gender, WHO grade, radiotherapy and chemotherapy status, and survival data were obtained for our further analysis. Similarly, the corresponding 1p19q codeletion and IDH mutation status were downloaded from CGGA database. Glioma samples were excluded who died for non-cancer related factors with survival times less than 30 days. In addition, a portion of glioma subjects with imperfect information were also eliminated. No specific ethical approval or patients informed consent was not required necessary because all of these data were publicly available.

### Identification of aging-related lncRNAs

The list of aging-related genes were acquired from Human Ageing Genomic Resources (HAGR; Table S1). To determine the correlation between aging-related genes and lncRNAs, we performed Pearson correlation coefficients in software R (version 3.6.3). The aging-related lncRNAs were selected out based on the criteria that correlation coefficient |R|> 0.6 and *P* value was less than 0.001. According to the above threshold values, candidated aging-related lncRNAs were identified and further analyzed.

### Cluster analysis of aging-related lncRNAs

We performed Principal Component Analysis (PCA) to visualize these aging-related lncRNAs expression patterns in different glioma patients in CGGA database and validated in TCGA database. The ConsensusClusterPlus algorithm was used for clustering samples assessments and then patients were divided into subtypes. The aging-related lncRNAs were identified three subtypes in CGGA.

### Development and validation of aging-related lncRNAs prognostic signature

To identify the potential prognostic-related lncRNAs, univariate Cox regression analysis was used to analyze the association between aging-related lncRNAs and overall survival (OS). The least absolute shrinkage and selection operator (LASSO) Cox regression was performed for filtrated more meaningful prognosis-related LncRNAs to establish the risk signature. Finally, we developed an aging-related lncRNAs prognostic signature for glioma patients involving fifteen aging-related lncRNAs. Then, we constructed the prognostic signature of aging-lncRNAs. According to risk coefficient and the expression levels of each lncRNA, and a risk score was calculated for each patient. The calculation formula of risk score is as follows: risk score= lncRNA1β × Expression + lncRNA2β × Expression + … lncRNAnβ × Expression.

### Prediction analysis of prognostic signature

On the basis of the median risk score as the threshold, the glioma patients were divided into high- and low-risk cohorts. We depicted survival curve between two cohorts using Kaplan-Meier curve method with a two-sided log-rank test. Univariate Cox regressions were utilized to evaluate the effect of clinicopathological variables on survival of glioma patients, including age, gender, tumor grade, radiotherapy and chemotherapy status. Furthermore, multivariate Cox regressions was performed to determine whether risk score was an independent prognostic factor. In addition, we performed stratified survival analysis to detect the impact of prognostic value of our risk scores model in different glioma subgroups. To further delve into the effect of single aging-related lncRNAs on glioma patients in the prognostic risk model, the relationship between expression of each screened lncRAN and clinical characteristics was assessed by Student’s *t*-test or one-way analysis of variance (ANOVA). In addition, predictive efficiency of the risk score was generated by calculating the area under the curve (AUC) of receiver operating characteristic (ROC) curve.

### Co-expression network and gene sets enrichment analysis

Pearson correlation coefficients were calculated among lncRNAs and aging genes in glioma patients using R software (v.3.6.3) (R > 0.6, *p* < 0.001). The aging-related lncRNAs and target genes co-expression network were constructed using Cytoscape (v.3.8.2). A Sandkey diagram was used to visualize the co-expression of mRNAs and LncRNAs and showed the risk type. To explore the Functional enrichment of the fifteen lncRNAs, we conducted gene set enrichment analysis (GSEA; v.4.0.3) to determine the biological functions and pathways by this priori defined lncRNAs and verified whether a showed statistically significant differences between high-risk and low-risk group or not.

### Immune infiltration analysis

We obtained immune cells infiltration score for each glioma patient to explore the degree of immune cells infiltration between low- and high-risk groups. And then we assessed the difference in proportion of 22 immune cell subtypes between the low- and high-risk groups. The ESTIMATE algorithm was applied to compare of the estimation, immune and stromal score between the two groups [a]. Furthermore, the scatter plot showed the correlations of risk score with Macrophages M0, Monocytes, and NK cells activated. The correlation between immune cells infiltration and risk scores was calculated by Pearson correlation (*P* < 0.05 was considered statistically significant).

### Somatic mutations analysis based on risk score

Further, we explored the genetic alteration of glioma patients in high- and low-risk groups and represent by waterfall plots. The top 10 somatic mutation were screened in two groups, respectively. In addition, according to different classifications, the mutations were further sorted in detail. The exclusive and co-occurrence of mutated genes in both two groups of glioma patients were visualized.

### Quantitative Real-Time Polymerase Chain Reaction (qRT-PCR)

We collected 7 glioma and non-neoplastic brain tissues (NBTs) tissues from patients who underwent surgical operation. Fresh tissue samples were frozen and stored at -80°C. This research was approved by Ethics Committee of Xiangya Hospital, Central South University. And the informed consent was acquired fro m involved patients. We extracted total RNA from tissues following manufacturer’s instructions. cDNA Synthesis Kit (TaKaRa) was utilized to reverse transcription. Then, qRT-PCR assay was performed following the reaction steps. The related expression of lncRNAs were normalized by GAPDH mRNA and calculated by 2^-^ΔΔCt method. Primers sequences were showed as follows: LINC00665 forward 5’ -GAGGACTCAGAGGTGGAATT-3’, and reverse 5’ -CAGCCAGC TTGTAGGG-3’; LINC00339 forward 5’ -ACCATGCTAGAAAGCCTCCC-3’,and reverse 5’ -CGTCCAGCAAGGTCCTAGAG-3’; SNHG16 forward 5’ -ACAT CGGCATGATGGCAGAA-3’, and reverse 5’ -TCACAAAAGGCGGGACCAC-3’; PAXIP1.AS2 forward 5’ -GCTTCCAGGGGAGAT-3’, and reverse 5’ -ATC AGACTGCCTGAAGA -3’; LINC00092 forward 5’ -CCTATGATTTGGCCTCT GGA-3’, and reverse 5’ -GAGAGCAGCGTTCAGGAAAC-3’.

### Statistical analysis

All statistical analyses were conducted using R software (v.4.0.5). Kaplan-Meier curves and log-rank tests were applied to evaluate survival data between various subgroups. Univariate and multivariate Cox regression analyses were used to assessed the independent prognostic factors for glioma patients. The risk coefficients of prognostic signature were calculated by LASSO regression. Prognostic accuracy of the nomogram and predicted signatures for 1-, 3-, and 5-year OS was estimated by ROC curves. Single nucleotide variation was analyzed with maftools R package. The differences between variables were evaluated by independent t-tests. Chi-square test was performed to predict the association of variables of clinical parameters between high- and low risk groups. Pearson and Spearman correlation analyses were conducted to determine the correlation. Probability value of less than 0.05 or 0.001 was regarded statistically significant.

## Results

### Identification of aging-related lncRNAs

We obtained 307 aging related genes from HAGR public database (Table S1). The RNA-sequencing profile data and corresponding clinical parameters of glioma patients were downloaded from CGGA and TCGA. A total of 928 lncRNAs were screened from CGGA and 629 lncRNAs were identified from TCGA (Table S2 and Table S3). Then, based on Pearson correlation analysis (| R | > 0.6 and *P* < 0.001), 226 and 152 aging-related lncRNAs were selected from CGGA and TCGA, respectively. We further performed univariate cox regression analysis to filtrate potential prognostic-related lncRNAs from aging-related lncRNAs, and found that 33 lncRNAs were significantly associated with OS of glioma patients. Detail data analyses were provided in Figure 1A. The correlation among 33 aging-related lncRNAs were presented by circle plot (Figure 1B).

**Figure 1.**
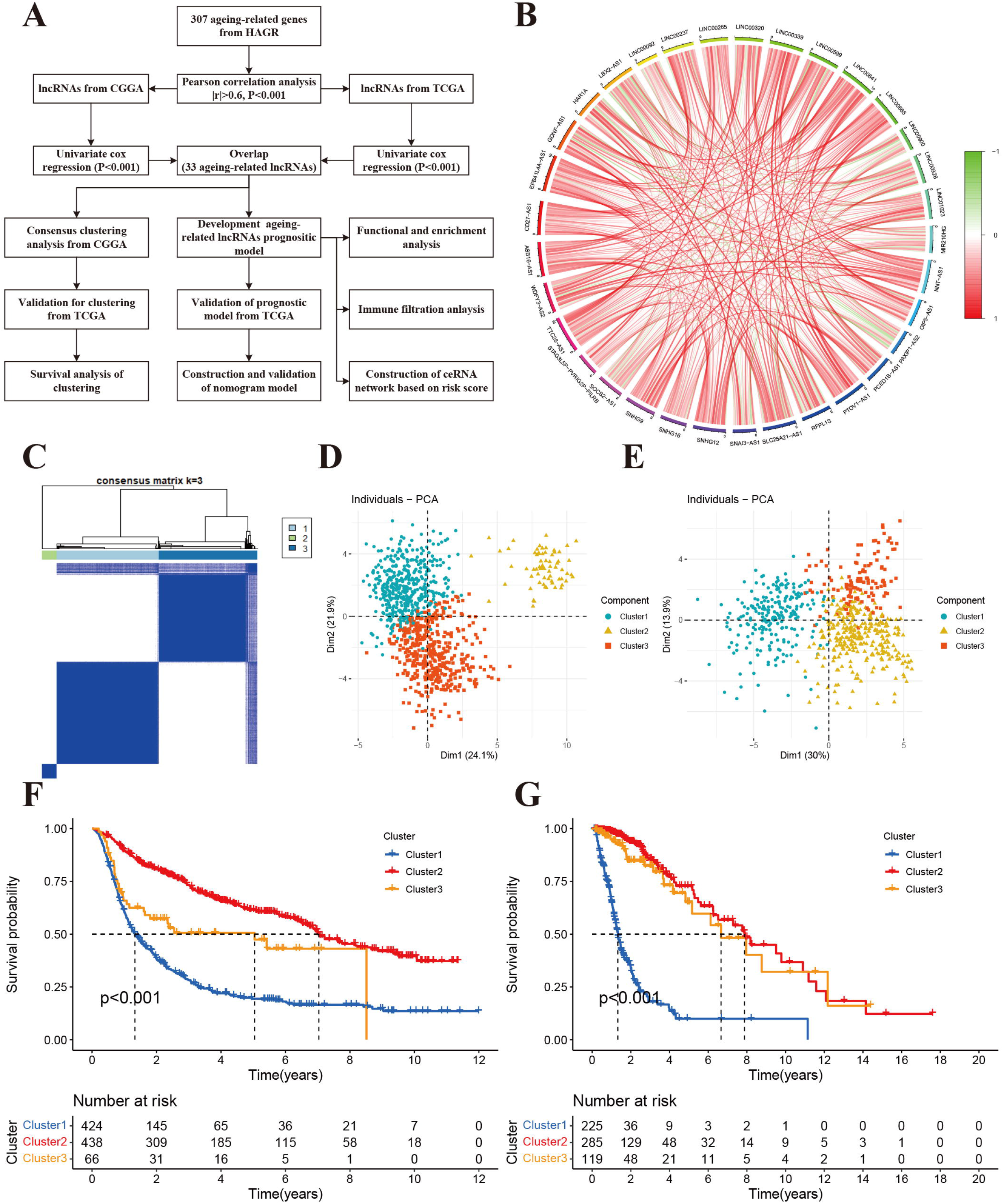
Molecular classification based on aging-related lncRNAs. **A:** The flow chart of data analyses in the study. **B:** The circle plot showed the correlation among 33 aging-related lncRNAs. **C:** Glioma patients were divided into three clusters in CGGA. **D:** PCA indicated that three subclasses were obtained in CGGA. **E:** Three subclasses were validated in TCGA. **F:** Kaplan-Meier cures of overall survival for three clusters in CGGA. **G:** Kaplan-Meier cures of overall survival for three clusters in TCGA.

### Clustering analysis of aging-related lncRNAs associated with prognosis

To identify aging-related glioma patterns, we performed ConsensusClusterPlus analysis to classify glioma patients into three subgroups using 33 lncRNAs (Figure 1C). The heat map showed sample clustering results with optimal clustering stability (the value of k=3). Subsequently, a PCA analysis suggested that glioma samples could be completely distinguished. There were three subtypes obtained in CGGA (Figure 1D) and validated in TCGA (Figure 1E). We further compared the prognosis of the three clusters by Kaplan-Meier cures of overall survival. In addition, the OS of the cluster1 was found to be shorter than these two clusters and the prognoses of the cluster2 were the best (Figure1F and 1G). The three clusters were observably separated in CGGA database (*P* < 0.001). While there is no different distribution between cluster2 and cluster3 about survival time in TCGA.

### Establishment and validation of aging-lncRNAs prognostic model

Based on the survival information of glioma samples, we applied univariate cox regression and screened 33 aging-related lncRNAs were highly related to OS. To better explore the prognostic role of those aging-related lncRNAs in glioma patients, we applied LASSO cox regression algorithm to validate that of 15 lncRNAs were most correlated with prognostics (Figure 2A). The optimal value of penalty parameter was determined for tuning the parameter selection in LASSO regression (Figure 2B). A risk score was calculated according to coefficients of each lncRNA (Figure 2C). The risk scores of glioma samples were calculated as follows: risk score = (0.220× Exp_LINC00665_) + (0.177× Exp_LINC00339_) + (0.126× Exp_SNHG16_) + (−0.026× Exp_PAXIP1.AS2_) + (0.035 × Exp_LINC00092_) + (0.030× Exp_LINC00265_) + (−0.164× Exp_SOCS2.AS1_) + (0.002× Exp_SNHG9_) + (−0.010× Exp_LINC00237_) + (−0.026× Exp_SLC25A21.AS1_) + (−0.078× Exp_EPB41L4A.AS1_) + (−0.126× Exp_HAR1A_) + (−0.164 × Exp_GDNF.AS1_) + (−0.262 ×Exp_SNA13.AS1_) + (−0.283 × Exp_WDFY3.AS2_). Taking the median risk score, glioma patients from CGGA and TCGA datasets were divided into high- and low-risk group, respectively. We found that there was a marked difference in prognosis between two groups (*P* < 0.001). Patients in high-risk group had a worse OS than than those in the low-risk group (Figure 2D, 2G). Further, the risk score and survival status distributions of glioma patients were showed in Figure 2E and 2H. Survival rates of glioma patients were correlated with risk scores, the mortality rate increased with a higher risk score. In addition, tSNE2 method were then performed to classify the samples into obvious two components in CGGA and TCGA (Figure 2F, 2I). Besides, ROC analysis indicated that risk score could accurately predict prognosis of glioma patients (1-year AUC = 0.760, 2-year AUC =0.832, 3-year OS =0.827 in CGGA and 1-year AUC = 0.858, 2-year AUC =0.889, 3-year OS = 0.9014 in TCGA; Figure 2J, 2K). These results demonstrated that the aging-lncRNAs prognostic model had a robust and stable prognostic value ability for glioma patients.

**Figure 2.**
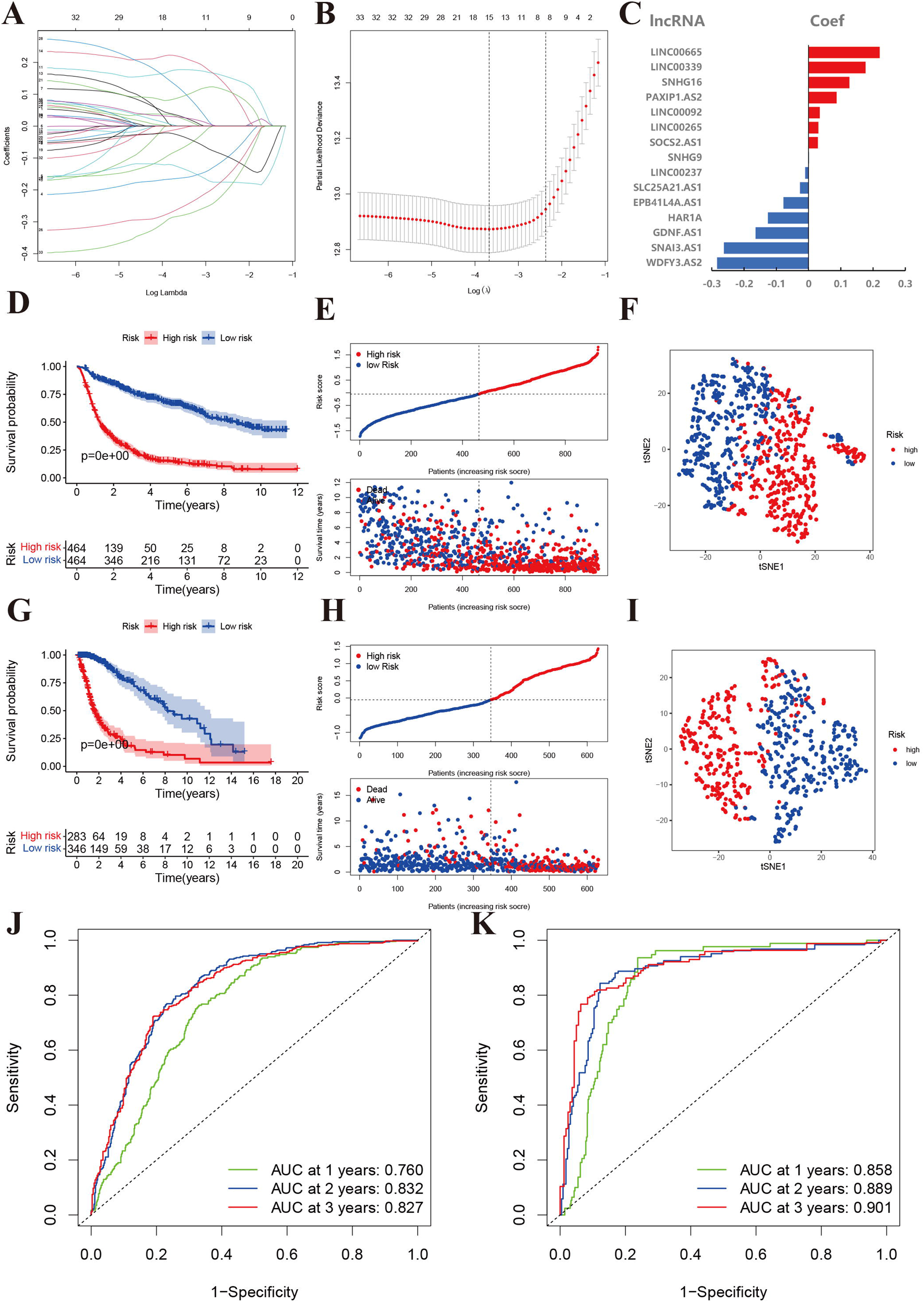
Development and validation of aging-related lncRNAs prognosis signature. **A:** LASSO regression of 15 aging-related lncRNAs. **B:** Cross-validation for tuning the parameter selection in the LASSO regression. **C:** Coefficient of prognosis model regression. **D:** Kaplan-Meier curves of high-risk group and low-risk group in CGGA. **E:** Distribution of risk score and patients based on the risk score in CGGA. **F:** The tSNE2 method showed obvious two components in CGGA. **G:** Kaplan-Meier curves of high-risk group and low-risk group in TCGA. **H:** Distribution of risk score and patients based on the risk score in TCGA. **I:** The tSNE2 method showed obvious two components in TCGA. **J and K:** ROC curves of prognostic signature based on risk score in CGGA and TCGA.

### Correlations of prognostic signature lncRNAs with clinical features

Finally, fifteen aging-related lncRNAs were involving in the prognosis signature. We performed univariate cox regression analysis to evaluate their prognostic roles. The forest plot illustrated that EPB41L4A.AS1, GDNF.AS1, HAR1A, LINC00237, SLC25A21.AS1, SNA13.AS1, and WDFY3.AS2 were protective factors with HR < 1, while LINC00092, LINC00265, LINC00339, LINC00665, PAXIP1.AS2, SNHG16, SNHG9 and SOCS2.AS1 were risk factors with HR>1 in glioma patients (Figure 3A). Subsequently, we investigated the distribution of clinic pathological features and expression of the fifteen aging-related lncRNAs between high- and low-risk groups, which for testing whether the aging-related lncRNAs signature could predict glioma clinical parameters. The heat map indicated that there were significant differences in age, grade, PRS type, histology, clusters, chemotherapy status, 1p19q codeletion and IDH mutation status (*P* < 0.01) between high- and low-risk groups (Figure 3B). Then, we further explored the relevance of risk score and each clinicopathological features. As expected, we found that the score of clusters 3, older age, advanced grade tumors, 1p19q non-codeletion, IDH wild type, recurrent tumors and chemotherapy status were significantly increased (Figure 3). The above results elucidate the aging-related lncRNAs signature may serve a pivotal role in tumor progression of glioma.

**Figure 3.**
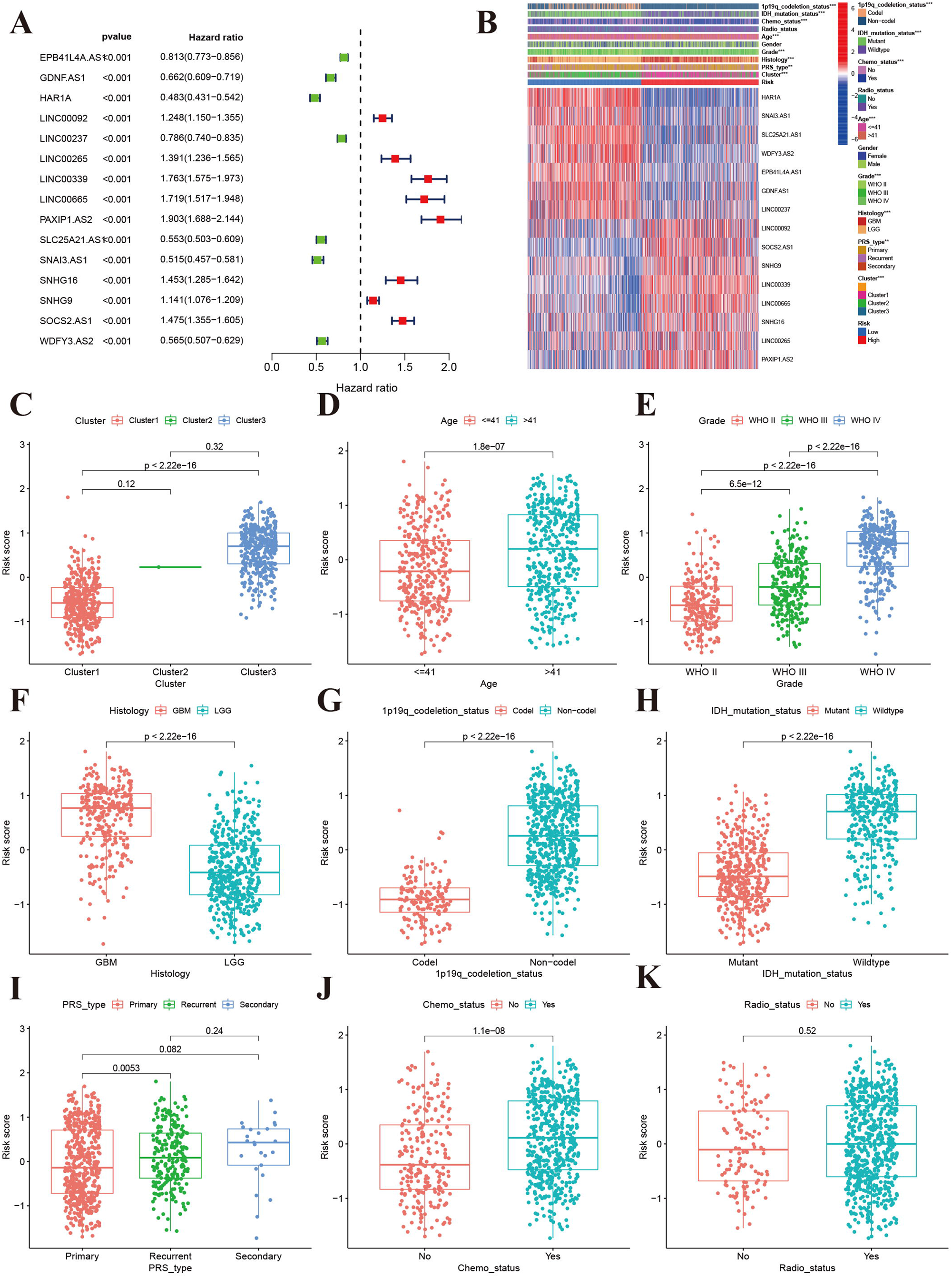
Correlations of clinical characteristic with identified aging-related lncRNAs signature. **A:** Forest of univariate COX regression for 15 signature lncRNAs. **B:** Heatmap showed that correlation of clinical parameters with risk scores and expression of 15 lncRNAs in high- and low-risk group. Boxplot showed the comparisons of risk score in different subgroups: **C:** Cluster1 vs Cluster2 vs Cluster3. **D:** age<=41 vs >41, **E:** WHO II vs WHO III vs WHOIV. **F:** GBM vs LGG, **G:** 1p19q_codeletion vs non-codel. **H:** IDH mutation vs wildtype. **I:** Primary vs Recurrent vs Secondary. **J:** Chemotherapy (Yes vs No). **K:** Radiotherapy (Yes vs No)

In addition, to prove the applicability of our prognostic model, we performed stratification analysis to identify whether it had ability to evaluate prognosis in each subgroup. As shown in Figure 4, in contrast with higher risk group, lower risk patients had longer survival time by Kaplan-Meier survival curve analysis. However, the OS rate was statistically similar for patients with secondary tumor, which because of the smaller sample size probably. These results indicated that our prognosis risk signature may be a potential predictor for glioma patients.

**Figure 4.**
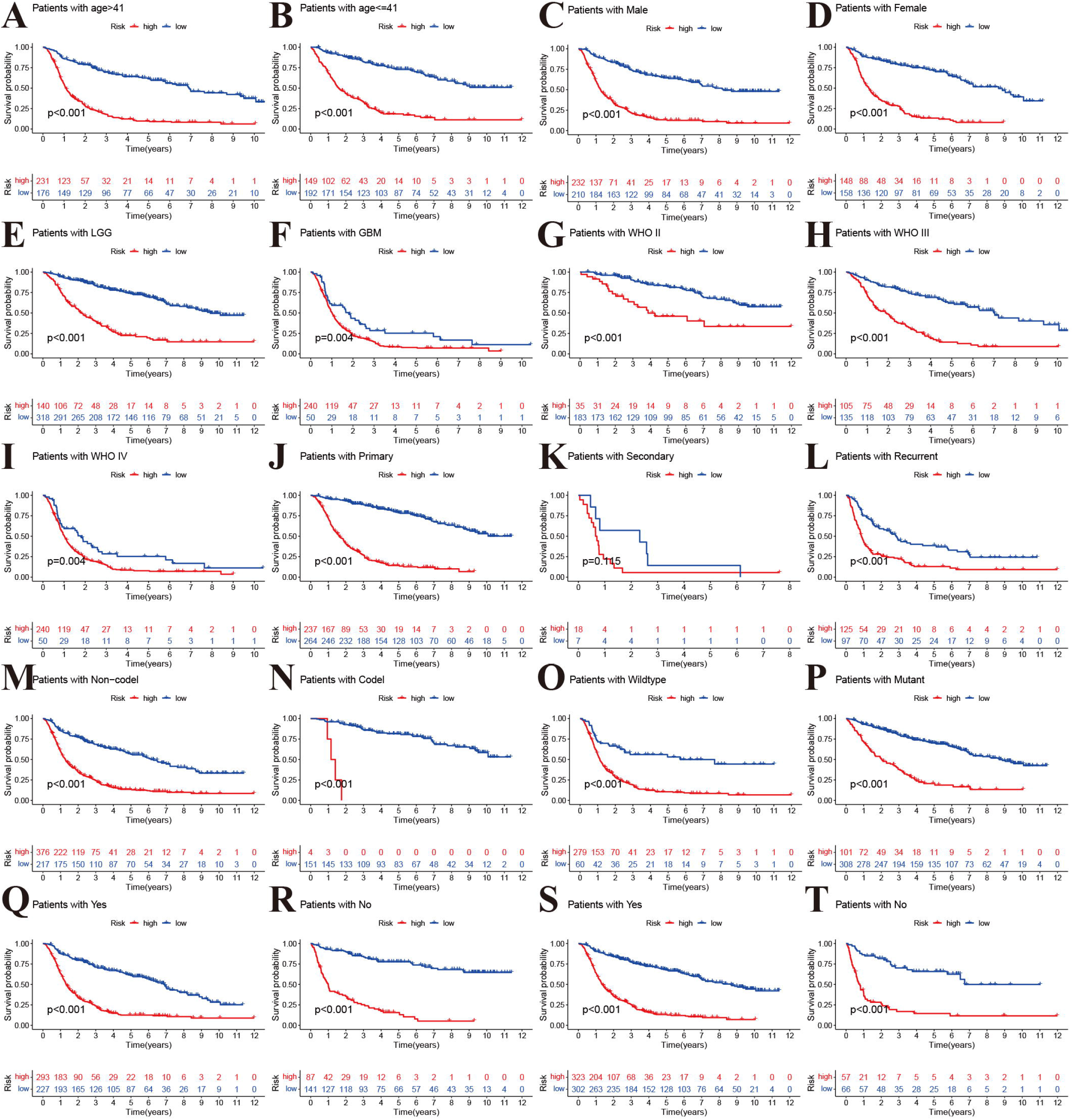
Stratified analyses of high- and low-risk group. **A and B:** age. **C and D:** Gender. **E and F:** Histology. **G-I:** WHO stage. **J-L:** Pathology type. **M and N:** 1p19q codeletion status. **O and P**: IDH mutation status. **Q and R:** Chemotherapy. **S and T:** Radiaotperapy.

### Prediction ability and independent analysis of prognostic signature

Univariate and multivariate Cox regression analysis indicated that age (HR = 5.296, 95% CI: 3.936–7.1263, *p* < 0.001), grade (HR = 4.662, 95% CI: 3.740–5.810, *p* <0.001) and risk score (HR = 6.556, 95% CI: 5.119–8.396, *p* < 0.001) were remarkably associated with OS in CGGA training cohort. Meanwhile, similar conclusions were observed in TCGA validating cohort (Figure5A, B). In order to develop a clinically applicable tool to predict OS in glioma patients by taking risk score of aging-related lncRNAs prognostic signature and other clinicopathological parameters. We built a nomogram to evaluate the probability of survival at 1, 3 and 5 years. The C-index value of the nomogram was 0.801(95%CI:0.783-0.820). Calibration curves demonstrated that concordances between actual and predicted of survival rates of glioma patients after bias corrections of nomogram in CGGA cohort (Figure 5C-E). These results indicated that our aging-related lncRNAs prognostic signature was reliable and precise.

**Figure 5.**
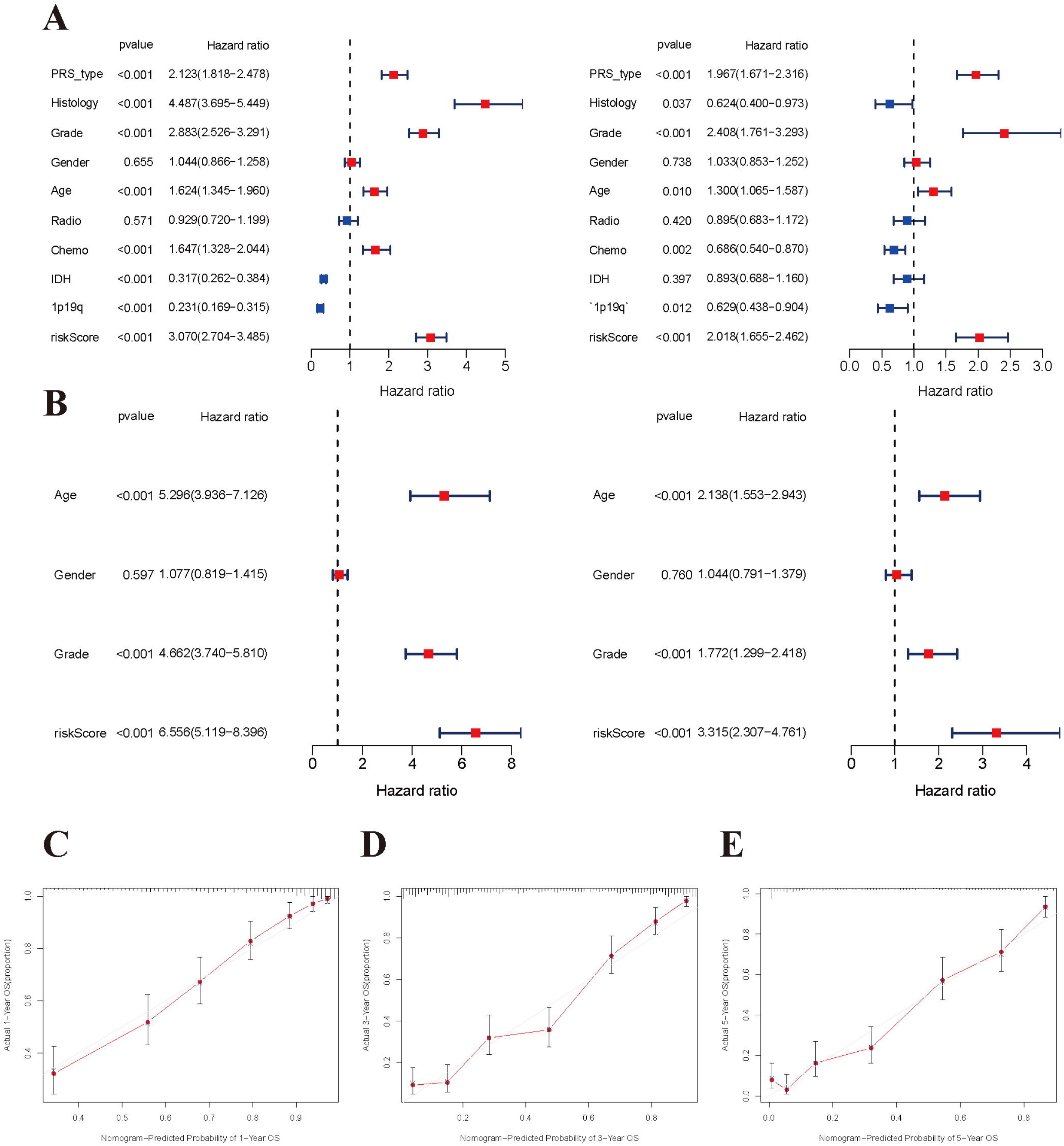
Independent prognosis analysis of risk score. **A and B:** Univariate and multivariate cox forest plot of risk score in CGGA and TCGA. **C-E:** Calibration plots of the nomogram for predicting the probability of OS at 1, 3, and 5 years in the CGGA.

**Figure 6.**
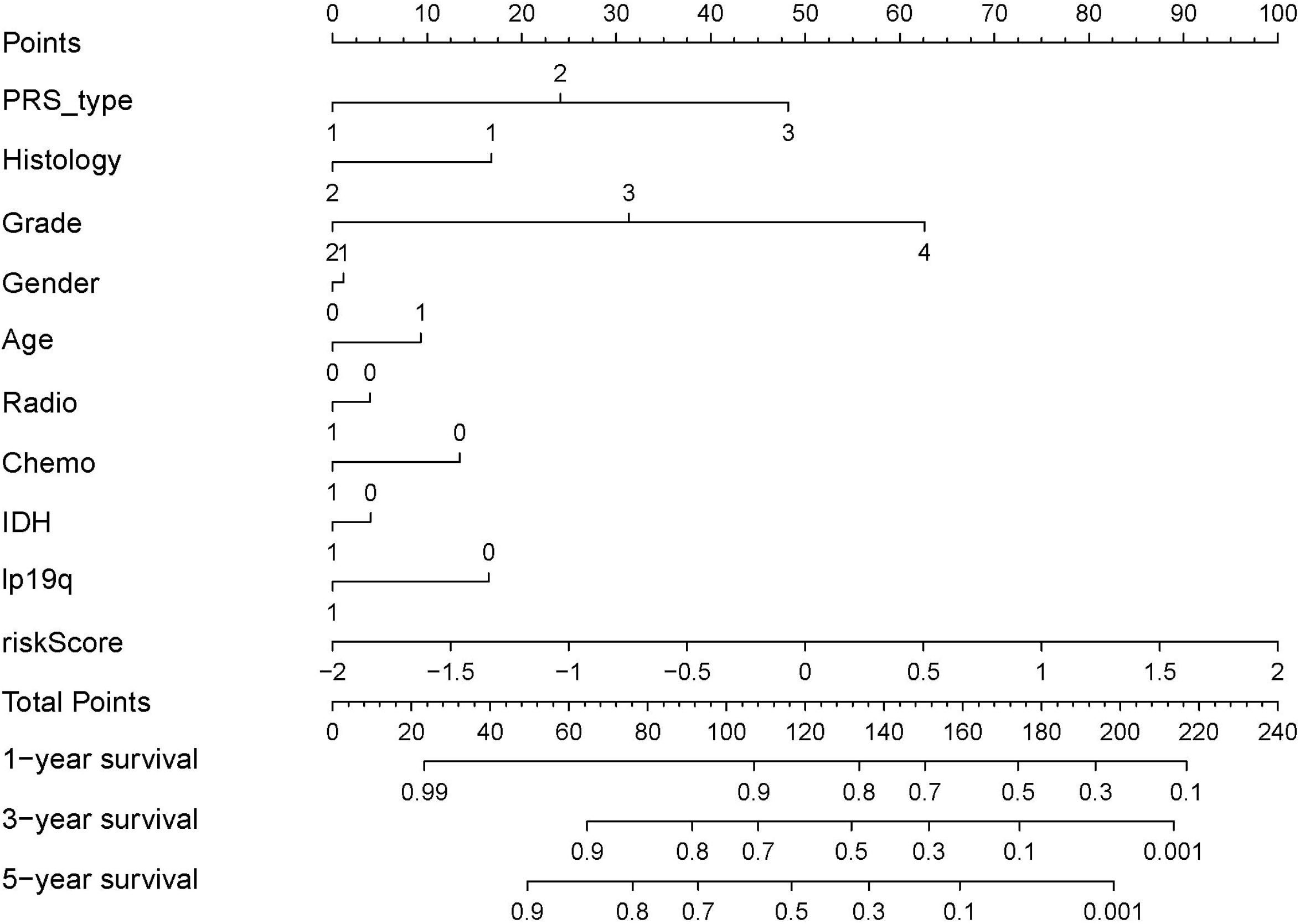
Nomograph of 1-, 3-, and 5-year overall survival probabilities predicted based on aging-related LncRNA signature.

### lncRNA-mRNA co-expression and pathways enrichment

Considering that miRNA and lncRNA can affect progression of cancers through mutual regulation, next we explored the potential functions of the fifteen aging-related lncRNAs in glioma by establishing a co-expression network. We found that fifteen target lncRNAs were closely correlated with 119 genes, which constructed a complex co-expression network. The detailed showed in Figure 7A). A Sankey diagram was depicted to visualize the relationship among lncRNAs, mRNAs and outcomes (risk/protective) (Figure 7B). In addition, KEGG analysis was performed to study the potential biological function and pathway in high- and low-risk groups. We observed that high-risk group was enriched in lysosome, N glycan biosynthesis, pathogenic Escherichia coli infection, primary immunodeficiency, primidine metabolism and regulation of actin cytoskeleton, and low-risk group was enriched in long term depression, long term potentiation, neuroactive ligand receptor, phosphatidylinositol signaling, and WNT signaling pathway, respectively. The top six significant gene sets in the two groups were presented in Figure 7C, 7D. The above data provided valuable insights for future finding potential individualized treatments in different risk score group of glioma patients.

**Figure 7.**
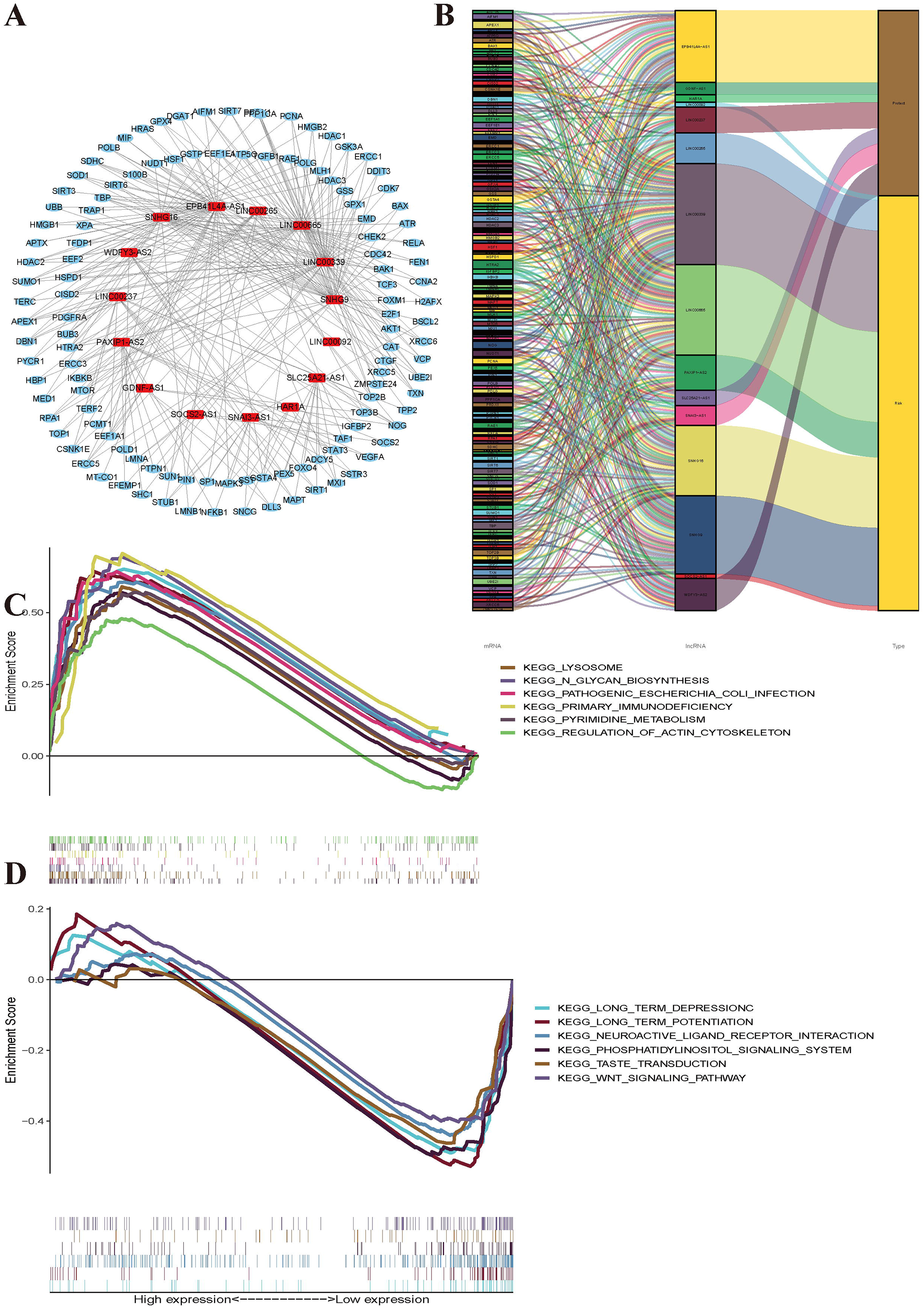
Functional and enrichment pathways analysis. **A:** LncRNAs-mRNA co-expression regulatory network based-on fifteen aging-related lncRNAs. **B:** A Sankey diagram was depicted to visualize the co-occurrences of lncRNAs, mRNAs and outcomes. **C:** KEGG pathway enrichment analysis in high-risk group. **D:** KEGG pathway enrichment analysis in low-risk group.

### Immune status based on risk score

In order to further investigate the various infiltration of immune cells between high- and low-groups, we compared the differences of 22 immune cells infiltration in the two groups. The proportions of B cells naive (*p*=0.030), plasma cells (*P*=0.046), T cells CD8 (*P*<0.001), T cells CD4 memory activated (*P*=0.002),T cells regulatory (*P*=0.001), T cells gamma delta (*P*=0.001=3), NK cells resting (*P*=0.021), Macrophages M1 (*P*<0.001), Macrophages M2 (*P*<0.001), Mast cells resting (*P*=0.005) and Neutrophils (*P*=0.004) were increased in high-risk group (Figure 8A). Besides, we calculated ESTIMATE score, immune score and stromal score by using ESTIMATE algorithm in two groups. Our result indicated that the scores of high risk group were higher than those of low risk group (*P*<2.22e-16) (Figure 8B-D). As shown in Figure 7F and 7G, risk scores were negatively correlated with the activation of Monocytes (R=058, *P*=2.2e-16) and NK cells (R=0.43, *P*=4.3e-10). However, the Macrophages M0 activation was upregulated with increased risk score (Figure 8E). These findings suggested that different immune infiltration status was emerged between two groups of glioma patients.

**Figure 8.**
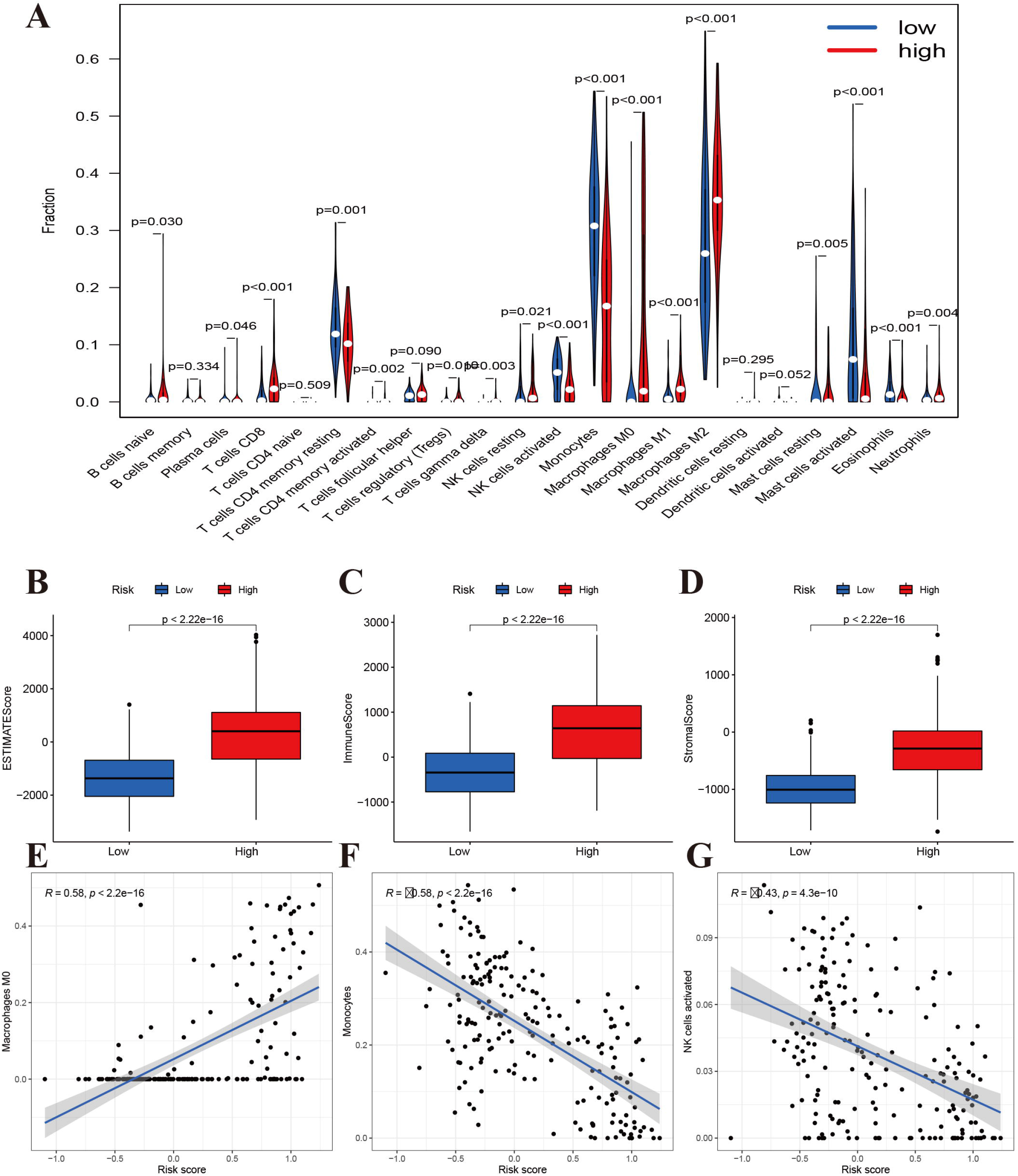
Immune filtration analysis between high- and low-risk groups. **A:** Differential analysis of immune-related cells based on risk score. **B-D:** Boxplot showed the comparisons of Estimation, immune and stromal score between high- and low-risk groups. **E-G:** Scatter plot showed that the correlations of risk score with Macrophages M0, Monocytes, and NK cells activated

### Somatic mutations analysis based on risk score

In addition, we performed somatic mutation profiles to analyze the gene mutation in high- and low-risk groups of 604 glioma patients. The waterfall plots exhibited that top 10 mutated genes were TP53, IDH1, EGFR, TTN, PTEN, ATRX, MUC16, FLG, PIK3CA and RYR2 in high-risk group, and were IDH1, TP53, ATRX, CIC, IDH2, PIK3CA, TTN, MUC16, 6MARCA4 and DNMT3A in low-risk group (Figure 9A,9B). As we can see, some mutated genes were high mutation frequency in two groups. Based on different classifications, mutations were further sorted. The missense mutations were accounted for the majority in two groups. And the most frequent variant types were single nucleotide polymorphism (SNP), of which occurred as C > T and T > C in low-risk group and C > T and C > A in high group (Fig, 9C,9D). Recently, co-occurrence and mutual exclusivity of genetics were often observed in cancer. In high-risk group, the co-occurrence mutations were much than those of in low-risk group (Details were showed in Figure 9E, 9F). However, they display significant exclusivity of mutations, such as IDH1 were mutually exclusive with PTEN and EGFR in high-risk group, while were mutually exclusive with IDH2 in low-risk group. This interrelated mutation could suggest functional interactions, which may provide new insights into clinical treatment.

**Figure 9.**
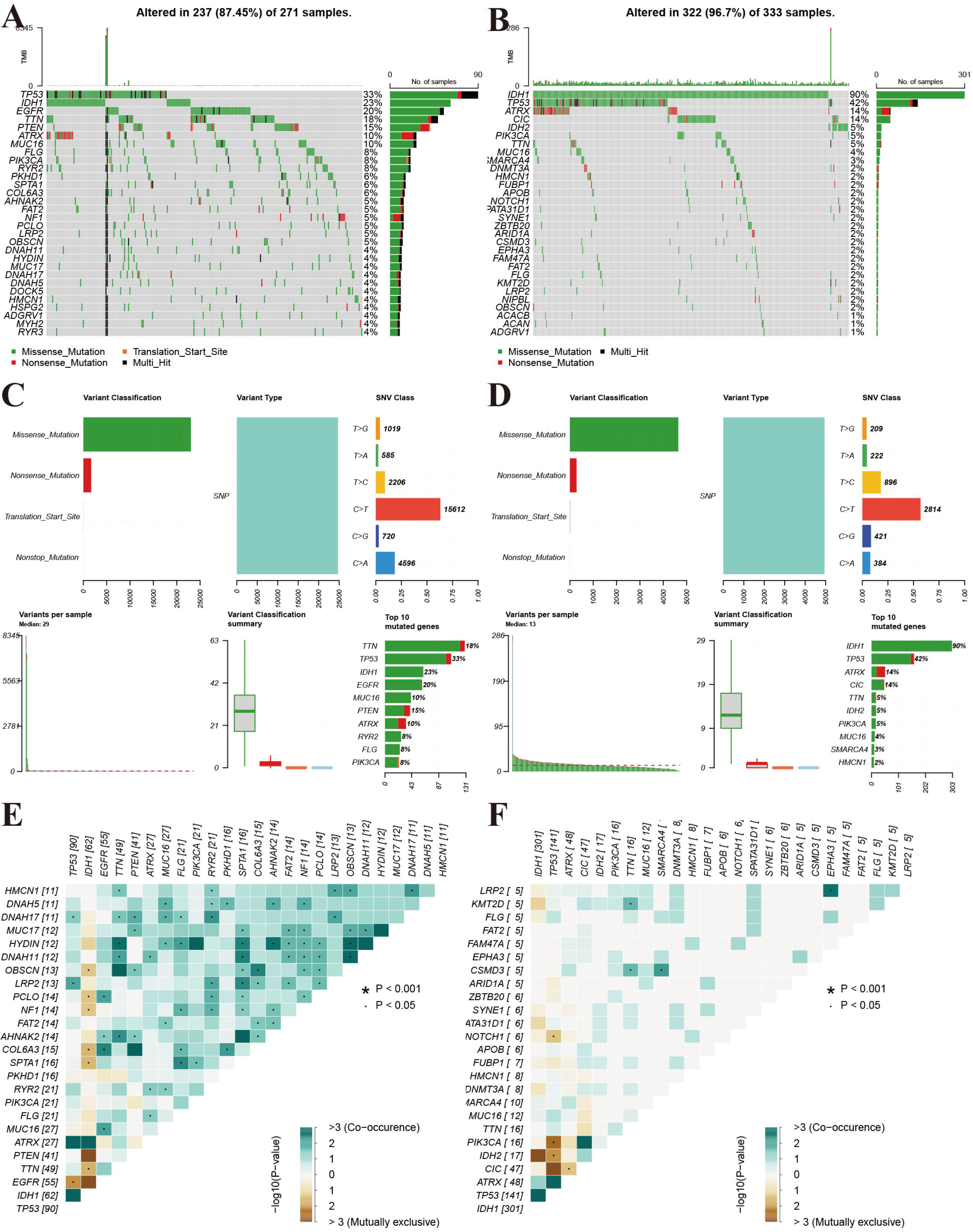
Landscape of mutation profiles between high- and low-risk glioma patients. **A and B:** Waterfall plots showed the mutations information in each sample of high- and low-risk group glioma patients. **C and D:** The variant classification in high-and the low-risk group glioma patients. **E and F:** The exclusive and co-occurrence in high-and the low-risk group glioma patients.

### Validation of aging-related lncRNAs expression levels

To further verify our study, we detected five aging-relate lncRNAs expression in 7 gliomas (4 WHO II gliomas and 3 WHO III gliomas) and NBT samples by qRT-PCR assay. Our results showed that mean expression levels of LINC00665, LINC00339, SNHG16, PAXIPI.AS2 and LINC00092 in glioma tissues were higher than those in non-tumor brain tissues ((Supplementary Figure S1A-E). Moreoer, the higher-grade patients were detected to obtain the higher lncRNAs expression. The above results confirmed the reliability of our analysis.

## Discussion

Glioma has a high recur and lead to a mortal outcome. The therapeutic effect of glioma is remains not satisfactory, especially glioblastoma. It is necessary for highly heterogeneous disease of glioma to identify potential prognostic indicators. Accumulating evidences provided that lncRNAs show a pivotal role in the process of tumor occurrence, development, metastasis, drug resistance and display the potential as a novel biomarker[17]. Moreover, with wide application of high-throughput technologies and the increasing maturity of data-sharing, tumor data about lncRNAs are accumulating in the public databases[18]. LncRNAs modulate diverse biological processes and the role in aging have recently attracted attention. It can regulate cell senescence, telomere length, and stem cell differentiation in aging process[19].

Aging is an inevitable process and is considered one of the predominant risk factors for most chronic diseases, including cancer[20]. Aging and cancer are interrelated. Aging-related genes can regulate cell senescence and tumor malignancy. The current view is that cell aging may promote occurrence and development of gliomas, because gliomas are more common in the elderly patients, who’s the number of senescent cells is increased dramatically in the brain[21]. Some scholars have also found that aging-associated genes linked to progression and prognosis of gliomas[22, 23]. Because of DNA damage from radiochemotherapy therapies have been shown to induce cells aging, which may be associated with glioma recurrence after treatment. Moreover, aging brain cells secrete excessive levels of factors to promote cell survival and invasion, such as MMP-2 and MMP-9[24, 25]. At the present, studies on indicators of glioma based on lncRNA signature is mounting. However, the potential role of aging-related lncRNA prognosis signature in glioma is not adequately researched. This study was performed to assess the role of aging-related lncRNAs in glioma using CGGA dataset as training cohort and TCGA dataset as validation cohort.

In this study, we identified a fifteen aging-related lncRNAs risk signature in gliomas through uni-cox regression and LASSO analysis. Finally, eight lncRNAs (LINC00092, LINC00265, LINC00339, LINC00665, PAXIP1.AS2, SNHG16, SNHG9 and\ SOCS2.AS1) were associated with high risk, and patients with high expression of these lncRNAs had an unfavorable prognosis. The remaining seven lncRNAs (EPB41L4A.AS1, GDNF.AS1, HAR1A, LINC00237, SLC25A21.AS1, SNA13.AS1, and WDFY3.AS2) were related to low risk. The AUC values to predict the OS of glioma in training cohort (1-year AUC = 0.760, 2-year AUC =0.832, 3-year OS =0.827 in CGGA and 1-year AUC = 0.858, 2-year AUC =0.889, 3-year OS = 0.9014), which indicate that the prognostic risk model is reliable and stable. Recently, a large number of studies have revealed the important role of lncRNA as oncogenes or cancer suppressor genes in various tumors. Studies have presented that lncRNAs play a complex regulatory role in the progress of tumor. Zhao et al. found that LINC00092-silenced cells presented obviously compromised metastatic potential and reduced invasive capacity in ovarian cancer, and which involved in glycolysis process[26]. Some scholars have revealed that overexpression of LINC00665 could reversed the ability of invasion and migration through encoding micropeptide in triple-negative breast tumor cells[27]. Meanwhile, Lu et al. showed that LINC00665 regulate stemness and EMT properties to promote gemcitabine resistance in gemcitabine resistance[28]. And in glioma cells, LINC00665 could also inhibit tumor progress via STAU1-mediated mRNA degradation[29]. LncRNA WDFY3-AS2 as a ceRNA to inhibit invasion properties, which is correlated with lymph node metastasis and TNM stage in oesophageal squamous cell carcinoma[30]. Wu et al. indicated that WDFY3-AS2 upregulated SDC4 expression to promotes cisplatin resistance in ovarian cancer. And si-WDFY3-AS2 should reduce invasion and migration of tumor cells[31].

A PCA based on aging-related lncRNAs showed three significantly different clustering patterns in glioma samples, and there was a difference OS among three clusters. Continuous inflammatory response led to cancer. In addition, it should be noted that the relationships between aging-related lncRNAs and immune cell infiltration. Our result revealed that different immune infiltration status was emerged between two groups of glioma patients. The risk scores were negatively correlated with the activation of Monocytes and NK cells, while the Macrophages M0 activation was upregulated with increased risk score. Further, we systemically analyzed the corresponding key signaling pathways of those lncNRNAs by KEGG. The majority of enriched pathways manifested the immunomodulatory functions, and the top five significantly enriched pathways involved in phosphatidylinositol signaling, and WNT signaling pathway. Somatic mutations are generally considered as tumor-initiating events[32]. Then, we also used waterfall plots to show the common mutate in glioma, that TP53 and IDH1 genes were both high mutation frequency in two groups. This fifteen lncRNAs risk signature not only effectively predicted the prognosis of gliomas but also reflected clinicopathological factors, such as grade, chemotherapy status, 1p19q codeletion and IDH mutation status, which have the potential to be a precise indicator for prognosis of gliomas.

In conclusion, we constructed an independent and robust prognostic signature using fifteen aging-related lncRNAs. The present risk signature in our study was based on transcriptome databases, so validation with some fundamental experiments is required. Next, further to clarify investigation of underlying mechanisms and to reveal how it regulates infiltrating immune cells in glioma. This study may be beneficial for clinicians to more accurately identify patients with high risk factors and improve prognosis of glioma.

## Data Availability

All data produced in the present study are available upon reasonable request to the authors

## Data availability

Original data can be obtained from Supplementary materials.

## Author contributions

ZZL designed this study and directed the research group in all aspects, including planning, execution, and analysis of the study. ZGY drafted the manuscript. YYL, and NL, collected the data. LZZ provided the statistical software, performed the data analysis, LZZ arranged the Figures and Tables. ZGY and SLF revised the manuscript. All authors have read and approved the final version of the manuscript.

## Supplementary materials

**Supplementary material 1**

**Table S1** Aging-related genes

**Table S2** 33 aging-related lncRNAs expression in TCGA

**Table S3** 33 aging-related lncRNAs expression in CGGA

**Supplementary material 2**

**Figure S1** Validations of several identified lncRNAs in glioma tissue. **A-E:** LINC00665, LINC00339, SNHG16, PAXIPI.AS2, LINC00092

