## Supplementary figures and images for "Functional implications of aging-related lncRNAs for predicting prognosis and immune status in glioma patients"

### Figure S1

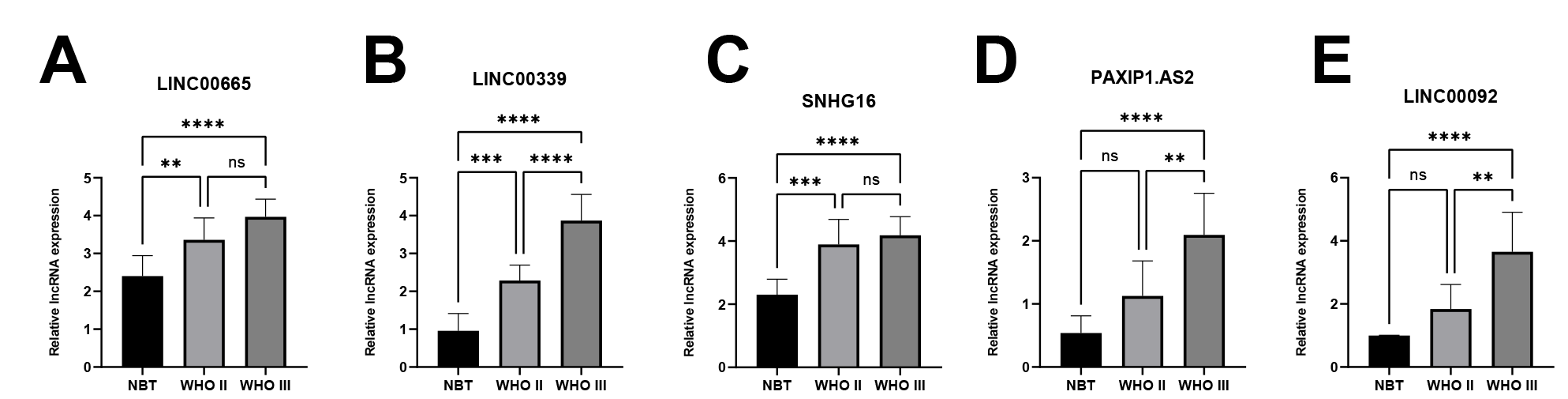
